# A prospective exposed-unexposed cohort study comparing time-to-pregnancy in patients with previous breast cancer and controls from a collaborative research network: results after three years of follow-up

**DOI:** 10.1101/2025.01.08.25320208

**Authors:** Anne-Sophie Hamy, Agathe Chabassier, Clara Sebbag, Christine Rousset-Jablonski, Clémentine Berkach, Isabelle Ray-Coquard, Laura Sablone, Lauren Darrigues, Elise Dumas, Angélique Bobrie, William Jacot, Marc Espié, Sylvie Giacchetti, Floriane Jochum, Aullène Toussaint, Geneviève Plu-Bureau, Lorraine Maitrot-Mantelet, Anne Gompel, Paul Gougis, Raphaëlle Bas, Christine Decanter, Bernard Asselain, Charles Coutant, Lili Sohn, Guillemette Jacob, Florence Coussy, Fabien Reyal, on the behalf of the Seintinelles Research Network

## Abstract

**Importance:** Data on fertility after breast cancer (BC) relative to the general population are lacking.

*Objective:* To compare the time-to-pregnancy between women with and without prior BC seeking to become pregnant.

*Design:* Prospective exposed-unexposed cohort study. Women were included from March 13, 2018 to June 27, 2019. Data were collected every six months via online questionnaires for up to three years.

*Setting:* Participants were recruited through the French collaborative network for cancer research Seintinelles*.

*Participants:* Exposed women (cases) were women aged 18-43 years with a history of localized BC who had completed treatment, without relapse. Unexposed women (controls) were women of the same age group with no history of BC.

**Exposure(s):** The exposure of interest was a prior history of BC.

**Main Outcome(s) and Measure(s):** The primary endpoint was time-to-pregnancy. The hypothesis tested was formulated before data collection. Secondary endpoints included the use of ART, including the use of cryopreserved material, factors associated with time-to-pregnancy, and pregnancy and neonatal outcomes. Statistical analysis was based on Kaplan-Meier survival analysis and multivariate Cox proportional-hazards models with adjustment for confounding factors by inverse probability of treatment weighting (IPTW).

*Results:* We enrolled 4351 women in this study. Follow-up data were available for 642 women (76 cases, 566 controls) who sought to become pregnant during the study period. Median time-to-pregnancy was 5 months (95% CI: 3 months to 7 months) for cases and 3 months (95% CI: 2 months to 5 months) for adjusted controls, with no significant difference between the groups (*p*=0.34). Cases were more likely to resort to the use of ART than controls (RR=13.9, 95% CI [2.2- 154.6]), but time-to-pregnancy was similar in cases and controls, with and without ART use. Time-to-pregnancy was influenced by factors such as age, parity, menstrual cycle regularity, BMI, and ART, but not by prior BC. Pregnancy and neonatal outcomes did not differ significantly between cases and controls.

**Conclusions and Relevance:** Time-to-pregnancy, in women seeking to become pregnant, was similar for women with and without a history of BC, raising hopes for women with BC wishing to have children.

## Introduction

Breast cancer (BC) is the most common cancer in women. There have been concerns for decades about the impact on potential recurrence of pregnancy after BC, but there are now large amounts of data suggesting that pregnancy after BC is safe ^1–4^.

Pregnancy rates in women with prior BC range from 3 to 13 %^4–7^, but these rates largely underestimate fertility, as patients who survive BC may not wish to become pregnant, and even if they do, they are confronted with several constraints on their plans: (i) it is recommended to delay pregnancy for at least 18 months after the end of treatment ^8^; (ii) pregnancy is contraindicated in patients treated with tamoxifen, due to its known teratogenic effects; (iii) the patient must have a partner and sexual intercourse, which may be difficult in up to two thirds of patients ^9,10^ and, finally, (iv) relapse may also compromise pregnancy plans. In addition, these women may have lower fertility due to the physiological decline in fertility if their pregnancy plans are delayed and/or due to chemotherapy-induced damage to the ovaries.

Many studies have investigated the relationship between chemotherapy and surrogate markers of fertility. Anti-Müllerian hormone (AMH) levels have been studied in detail, with the following findings: (i) AMH concentration is lower in BC survivors than in controls ^11–14;^ (ii) AMH concentration is lower after chemotherapy than before treatment ^11,12,15–18;^ (iii) After chemotherapy for BC, AMH is higher in menstruating women than in patients who are amenorrheic ^14,15,19^; (iv) AMH concentration before treatment is predictive of ovarian function, as it is based on menstrual function^11,16,17^. However, in most of these studies, ovarian reserve evaluations were not linked to reproductive outcomes and pregnancy. Several teams have shown that AMH levels increase again after chemotherapy ^20–22^, suggesting that this parameter is of little value as a dynamic marker of ovarian reserve. Furthermore, several studies have reported spontaneous pregnancies in women without detectable levels of AMH^20,21^. Evaluations of fertility loss after cancer therefore remain the Achilles’ heel of the oncofertility domain.

Published findings concerning the desire for pregnancy and success rates for conception after BC are encouraging. Partridge *et al.* ^23^ surveyed 440 women and found that 57% of those who attempted to become pregnant were successful. In an American prospective cohort study, 69.2% patients with a history of BC wishing to become pregnant conceived successfully ^24^. In the recent POSITIVE study of 497 patients with previous BC interrupting endocrine therapy to attempt to become pregnant for whom pregnancy status data were available ^25^, 74.0% of the patients reported falling pregnant at least once during the trial. However, pregnancy rates after BC have never been benchmarked against that in the general population, leaving questions about the theoretical effects of BC treatment on fertility unanswered. The primary objective of the FEERIC study was to compare time-to-pregnancy between women with and without prior BC seeking to become pregnant during a three-year follow-up period.

## Materials and methods

### Source of participants

Seintinelles* is a collaborative social network created in 2012 to accelerate the recruitment of French volunteers for cancer research studies, by connecting researchers with men and women of various ages, social and medical backgrounds with or without a history of cancer. In December 2024, the network included more than 40 000 French citizens willing to participate in research studies.

### Study design and inclusion criteria

The design of this study has been described extensively elsewhere ^26^. The FEERIC (Fertility, Pregnancy, Contraception after BC in France) study is a prospective case-control exposed-unexposed aiming to assess the impact of BC on fertility, pregnancy, and contraception in young BC survivors. Women were recruited from March 13, 2018, to June 27, 2019. Eligible exposed women (cases) were female patients aged from 18 to 43 years with a prior diagnosis of localized, BC (invasive or *in situ*) who had completed treatment (surgery, chemotherapy, and radiotherapy) at the time of enrollment and were relapse-free. Women who were under active endocrine therapy at the time of eligibility could also be included in the study and could decide to temporarily stop it when they sought to get pregnant, consistent with clinical practices. The exclusion criteria were prior hysterectomy and/or bilateral oophorectomy and/or bilateral salpingectomy. Eligible unexposed women (controls) were women aged from 18 to 43 years, free from BC and other cancers, who had not undergone hysterectomy, bilateral oophorectomy or bilateral salpingectomy. Volunteers matching the inclusion criteria were asked to complete a baseline form at inclusion and follow-up forms every six months for three years. Data were collected via self-administered online questionnaires released through the Seintinelles research platform. The Seintinelles scientific board approved the FEERIC project on December 7, 2015, and the Sud Ouest Outre Mer II ethics committee approved the project on October 5, 2017 (reference number: 2017: A0218152). As the target number of participants (*n*=1004) was reached prematurely, authorizations were obtained to pursue participant enrollment until the expected end of the study. The study follows the Strengthening the Reporting of Observational Studies in Epidemiology (STROBE) reporting guideline for case-control studies.

### Fertility and pregnancy endpoints

All fertility and pregnancy endpoints were self-declared. For each form, patients were asked to provide information about the following fertility and pregnancy endpoints: desire for a pregnancy (yes/no/not known); attempts to become pregnant (defined as current unprotected sexual intercourse with the intent of becoming pregnant); pregnancy outcomes.

***Time-to-pregnancy*** was defined as the time between the start of the attempt to become pregnant and the start of a pregnancy, regardless of the outcome of the pregnancy concerned. ***Assisted reproductive technology (ART***) was defined as medical procedures used primarily to address infertility issues, including treatments such as *in vitro* fertilization (IVF), intrauterine insemination (IUI), and other techniques facilitating conception, such as the use of embryos/oocytes/ovarian cortex cryopreserved before BC treatment. Women who conceived spontaneously were censored at the time of conception. In addition, as in the main analysis for time-to-pregnancy, women who stopped trying to get pregnant for personal reasons, experienced a disease relapse, or did not respond to the questionnaires were censored at the date of their last known follow-up.

***Time-to-ART*** was defined as the time from the start of the attempt to become pregnant to the first use of an ART method. The event of interest for these analyses was the first use of any ART method during an attempt to become pregnant beginning after inclusion in the study. Pregnancy outcomes were binned into three categories: full-term pregnancy, miscarriages and other abortive pregnancies (elective abortion, abortion for medical reasons or ectopic pregnancy, and other pregnancy outcomes). Deliveries were classified according to the route (vaginal or cesarean delivery) and the presence or absence of complications and fetal malformations.

### Study objectives

The main objective of the analysis was to compare the time-to-pregnancy between cases and controls, for women attempting to become pregnant during the three-year follow-up period of the study.

Other objectives included the evaluation of factors associated with time-to-pregnancy, and the comparison of the use of artificial reproductive technology (ART) and/or cryopreserved material reuse, obstetrical and neonatal outcomes between cases and controls (see the supplemental materials and methods).

### Statistical analysis

Standard descriptive statistics methods were used, as described in the supplemental material. We performed Kaplan-Meier survival analysis to evaluate potential differences in time-to-pregnancy from the start of the attempt to become pregnant until successful conception or censorship between cases and controls (see supplemental material and methods and Fig. S1). We ensured that the data points were independent by considering only the first recorded attempt to become pregnant for each woman.

We accounted for potential confounding factors (see supplemental material and methods and Fig. S2) by applying inverse probability of treatment weighting (IPTW) to adjust the Kaplan-Meier conception curves, bar charts and comparative analyses ^27^. We calculated ATT (average treatment on the treated) weights, also known as the standardized mortality ratio (SMR) in epidemiology ^28–30^. We used a propensity score estimated with a generalized linear model (GLM) ^31–33^ (see supplemental material).

We used a Cox multivariable proportional hazards model to evaluate the quantitative influence of various variables on time-to-pregnancy, including the impact of BC treatments ^34^. All statistical analyses were performed in R, with an α risk of 0.05 for hypothesis testing.

## Results

### Study participants

From December 2018 through June 2019, 4351 women were enrolled (cases *n*=517, controls *n*=3834) (Fig.1). In total, 1268 women withdrew from the Seintinelles network or did not complete any of the follow-up forms after inclusion in the study and were excluded from this analysis. There was a low gradual decrease in the number of respondents over the three-year study period (Fig. S3). On inclusion in the study, 2853 (cases *n*=367, controls *n*=2486) had available follow-up data and were not attempting to get pregnant. In total, 642 of these women (cases *n*=76, controls *n*=566, corresponding to 20.8% of responders), began trying to become pregnant during the time frame of the study and were included in the time-to-pregnancy analysis.

**Fig. 1:**
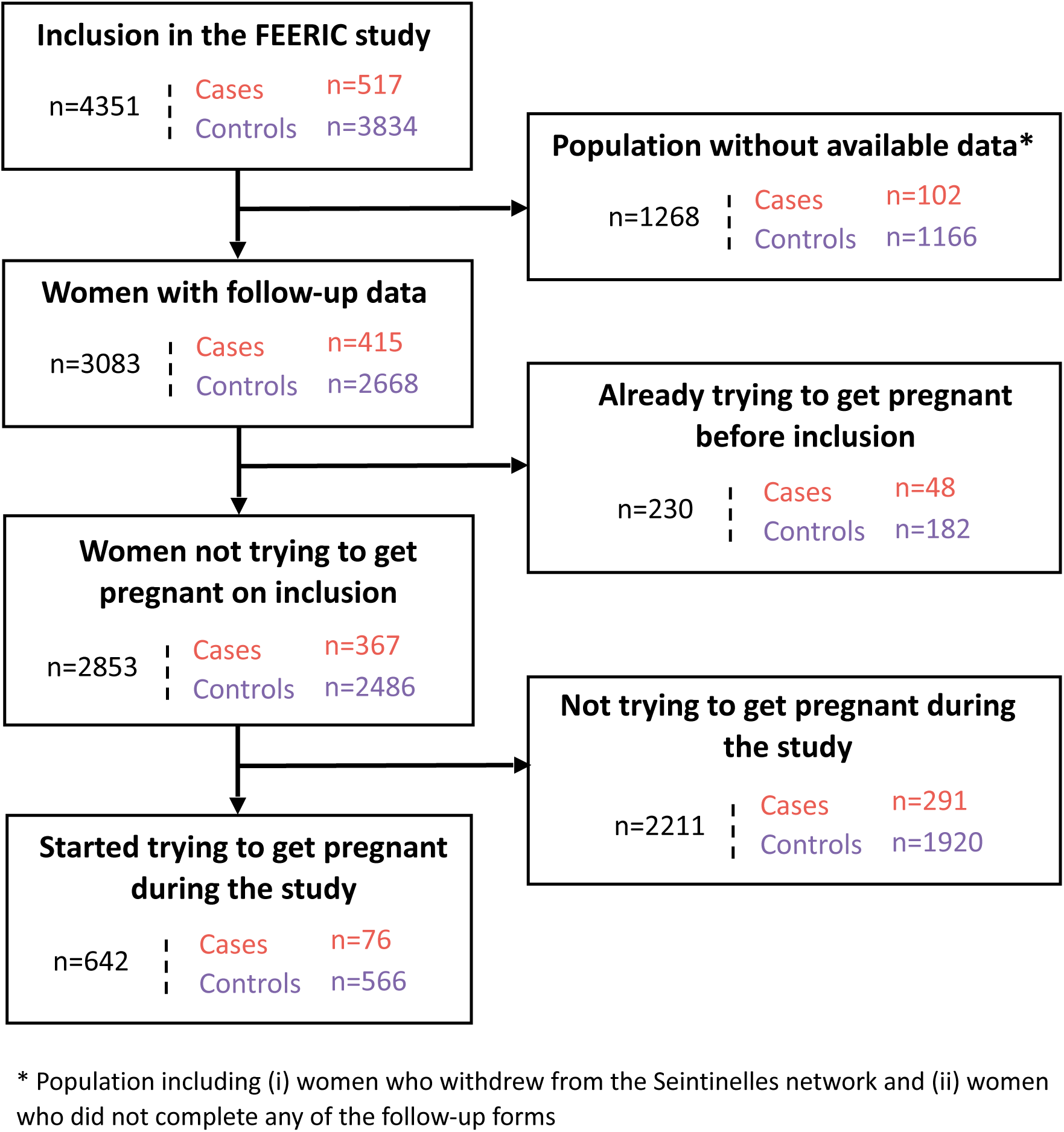
Flow chart for the study cohort.

The characteristics of the cases and controls were significantly different before adjustment. Cases were significantly older than controls at the inclusion in the study (35.0 versus 30.0 years old, *p*<0.001) (Table S1). Controls were made comparable to cases by implementing IPTW for 14 key factors in the control group (Fig. S4) making it possible to assess the causal impact of prior BC on fertility directly (Table1). After adjustment, age at start trying to get pregnant was 35.0 in cases and 35.7 in controls (p=0.59), most women had no children (cases 56.6% *versus* controls 54.5%, *p*=0.13). Most cases had prior chemotherapy (82.9%) or endocrine therapy (56.6%), and 59.2% had a fertility preservation procedure before treatment (oocytes 39.5%; embryos 15.8% ; and/or ovarian cryopreservation 4%).

**Table 1:**
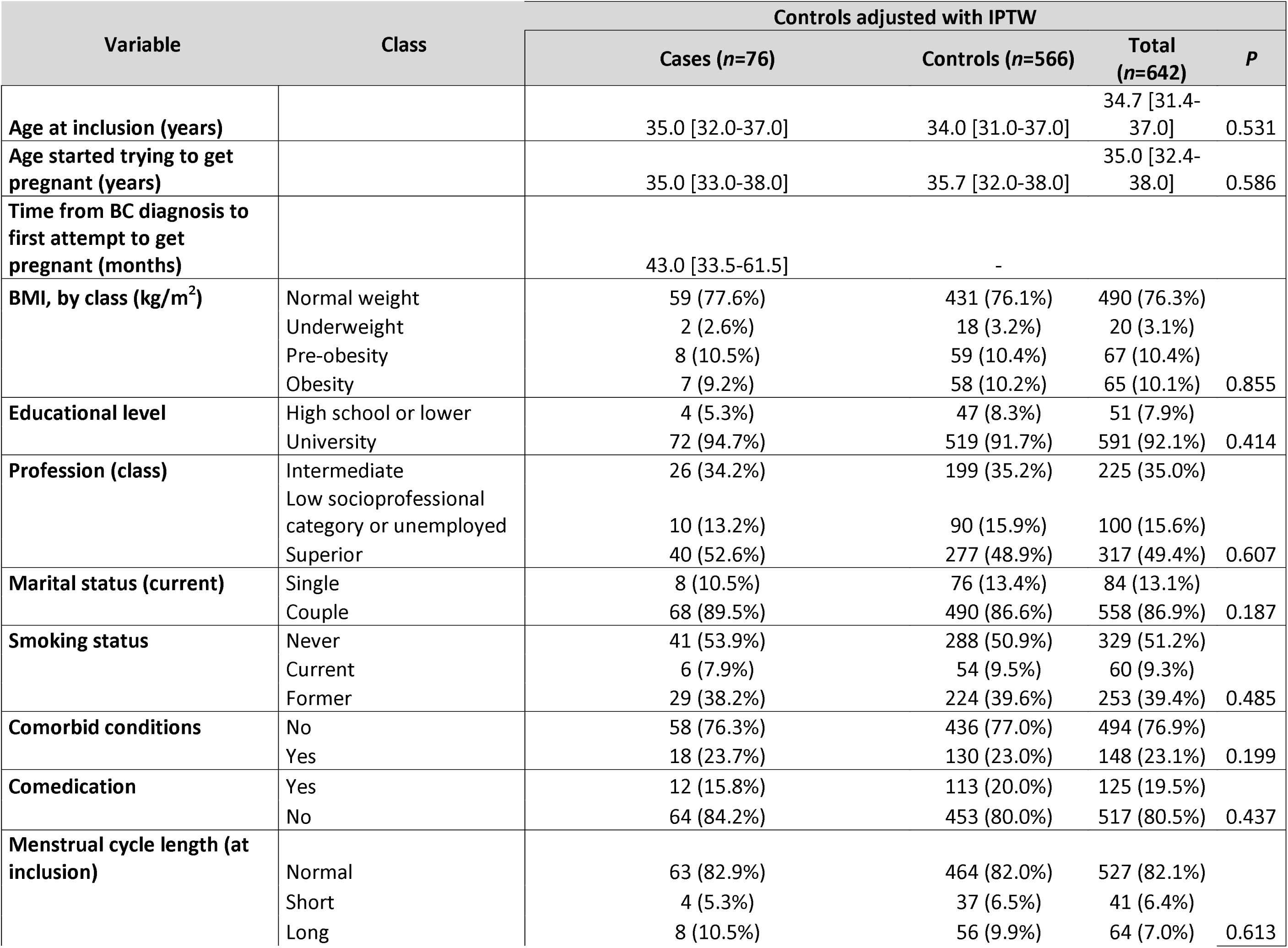
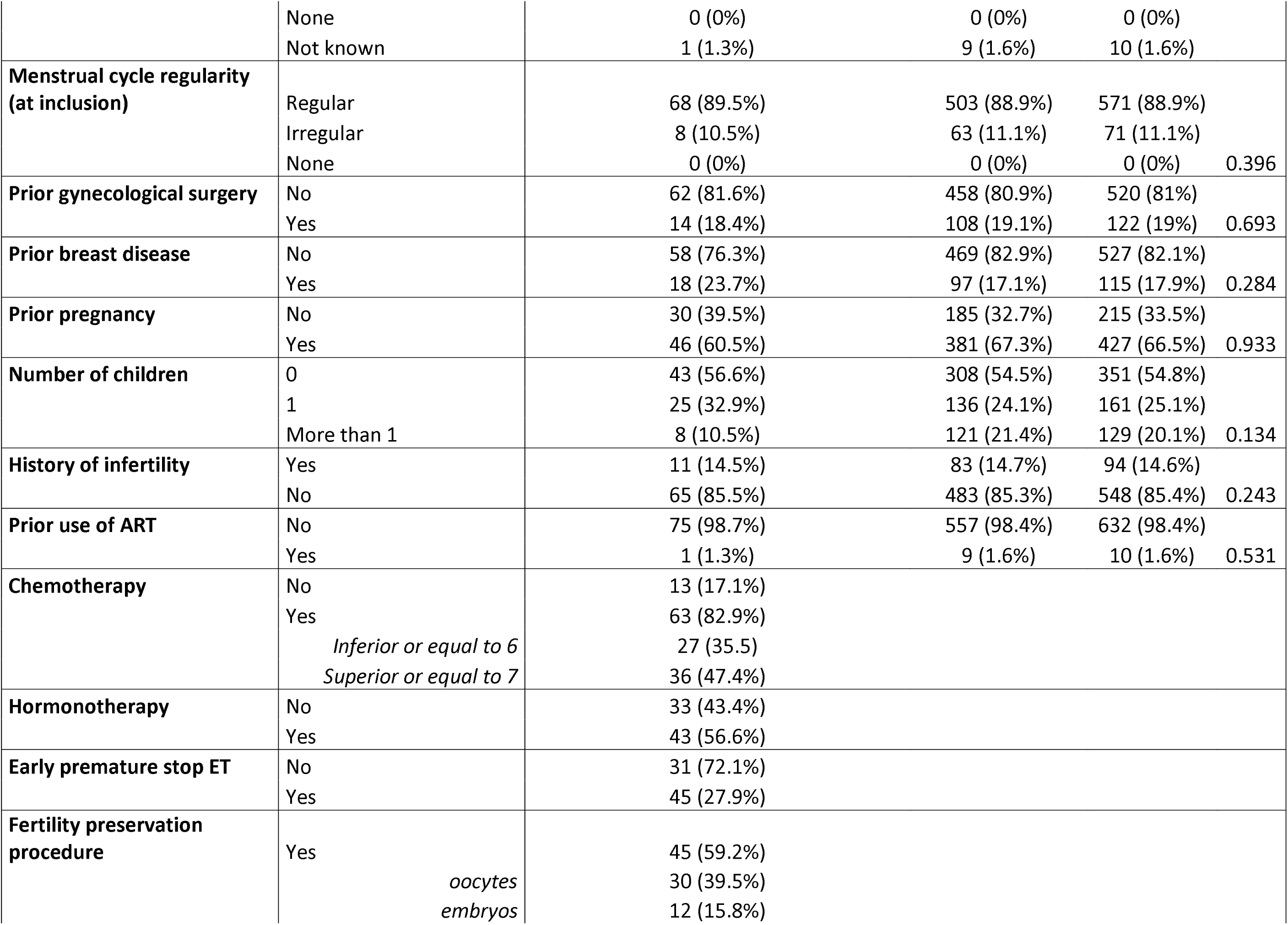
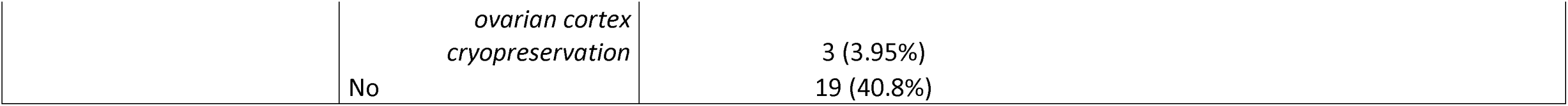
Characteristics of the study participants after inverse probability of treatment weighting (IPTW). This table compares various characteristics between breast cancer cases (*n*=76) and controls (*n*=566) adjusted with IPTW ATT weights. IPTW adjustment matches the distributions of confounding variables of the control This table compares various characteristics between breast cancer cases (*n*=76) and controls (*n*=566) adjusted with IPTW ATT weights. IPTW adjustment matches the distributions of confounding variables of the control and case groups. The confounding variables included age at which the woman first started trying to get pregnant, nulliparity, professional and educational levels, marital status, BMI, smoking status, infertility history, previous use ART methods, menstrual cycle length and regularity, prior gynecological surgery, prior breast diseases (before cancer for cases) and comorbid conditions. Proportions were calculated by applying IPTW ATT weights to the control group and the raw numbers were then determined by taking the closest integer to the product of the weighted proportion and the actual number in each category. *P*-values comparing distributions of variables between cases and controls demonstrate that significant differences existed between cases and controls for some variables before IPTW adjustment, but that these differences were no longer significant after adjustment, as no *p*-values were below the threshold after adjustment. Abbreviations: IPTW: inverse probability of treatment weighting, ATT: average treatment on the treated.

### Comparison of time-to-pregnancy between cases and controls

In the population of 76 cases and 566 weighted controls who attempted to get pregnant after inclusion, 50 cases (65.8%) and 402 controls (71.0%) became pregnant at least once. The median time-to-pregnancy was 5 months (95% CI [3.0-7.0 months]) for cases and 3 months (95% CI [2.0-5.0 months]) for controls and the distribution did not differ significantly between the two groups (*p*=0.34) (Fig. 2). One year after the women started trying to get pregnant, the cumulative incidence of pregnancy was 72.7% (95% CI, 59.8 to 84.3%) for cases and 78.5% (95% CI, 65.7 to 89.1%) for controls. Two years after the women started trying to get pregnant, these incidences had increased to 83.6% (95% CI, 68.4 to 100.0%) for cases and 84.2% (95% CI, 77.0 to 96.6%) for controls.

**Fig. 2:**
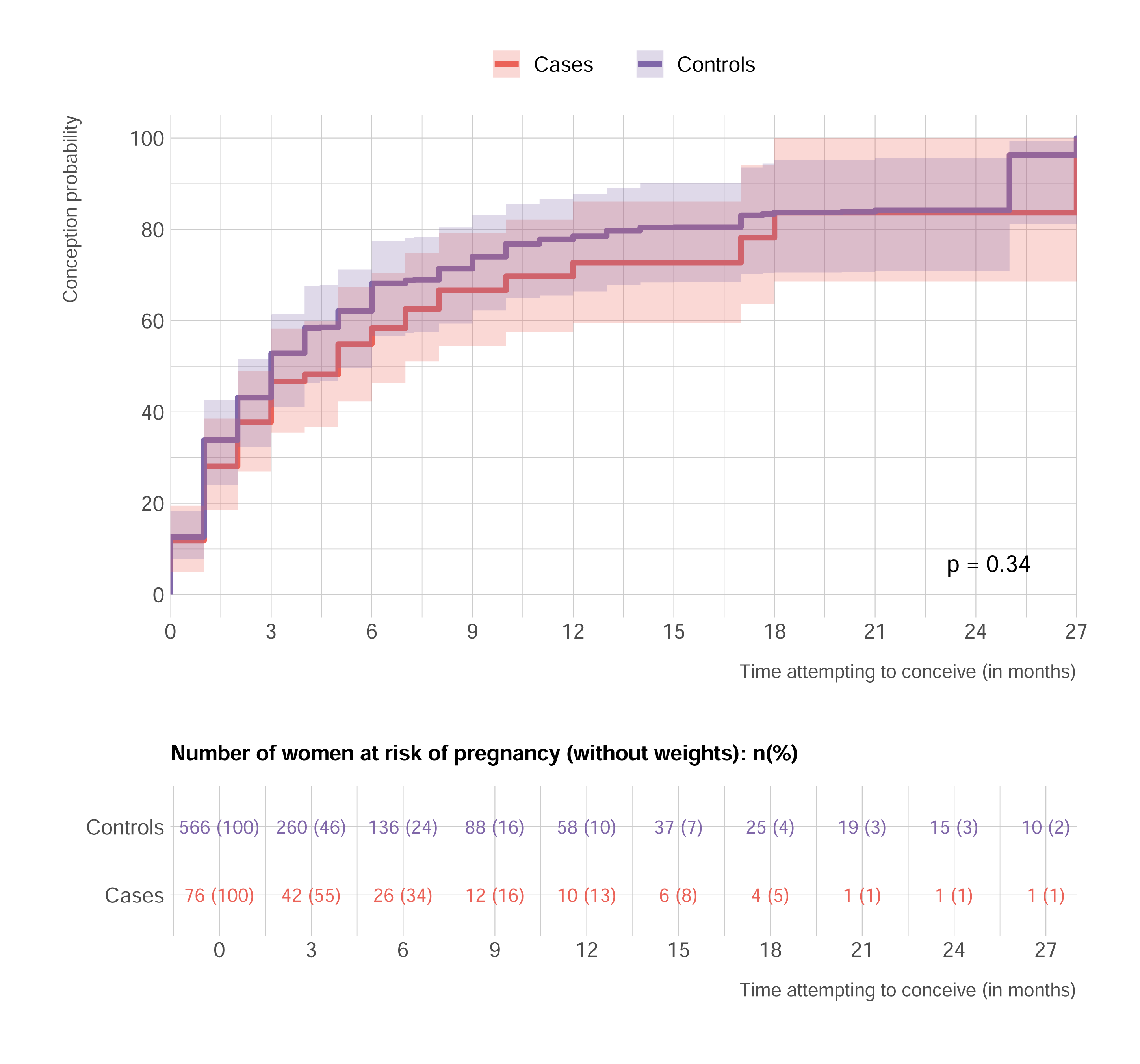
Time-to-pregnancy comparison between cases and controls. These curves depict the proportion of women attempting to get pregnant who became pregnant over time, with confidence intervals derived from 1000 bootstrap samples. The curves were adjusted by IPTW based on the calculation of ATT weights, to ensure a fair comparison. This approach balanced the distribution of 14 confounding variables between cases and controls, making the weighted control group comparable to the cases. Most women who attempted to get pregnant succeeded during the study period. In total, 19 cases and 111 controls were censored due to a lack of response or because they did not become pregnant before the end of the study. An additional 7 cases and 38 controls were censored because they changed their minds about trying to become pregnant. None of the 76 patients with prior BC who started trying to get pregnant after inclusion in the study experienced a relapse. Abbreviations: IPTW: inverse probability of treatment weighting, ATT: average treatment on the treated.

### Modes of conception and use of ART or cryopreserved material

Overall, 15 cases (19.8%) used either ART methods (*n*=7, 9.2%) or cryopreserved material (*n*=8, 10.5%; embryo, *n*=4; oocyte, *n*=3; ovarian cortex, *n*=1) in their attempts to get pregnant, and 25 weighted controls (4.4%) used ART. One year after the women started trying to get pregnant, the cumulative incidence of ART use was 41.1 % [17.8 – 62.6%] in cases *versus* 3.0 % [0.4-8.2] in controls, RR= 13.9 (95% CI: [2.2-154.6], *p*<0.001, Fig. 3).

**Fig. 3:**
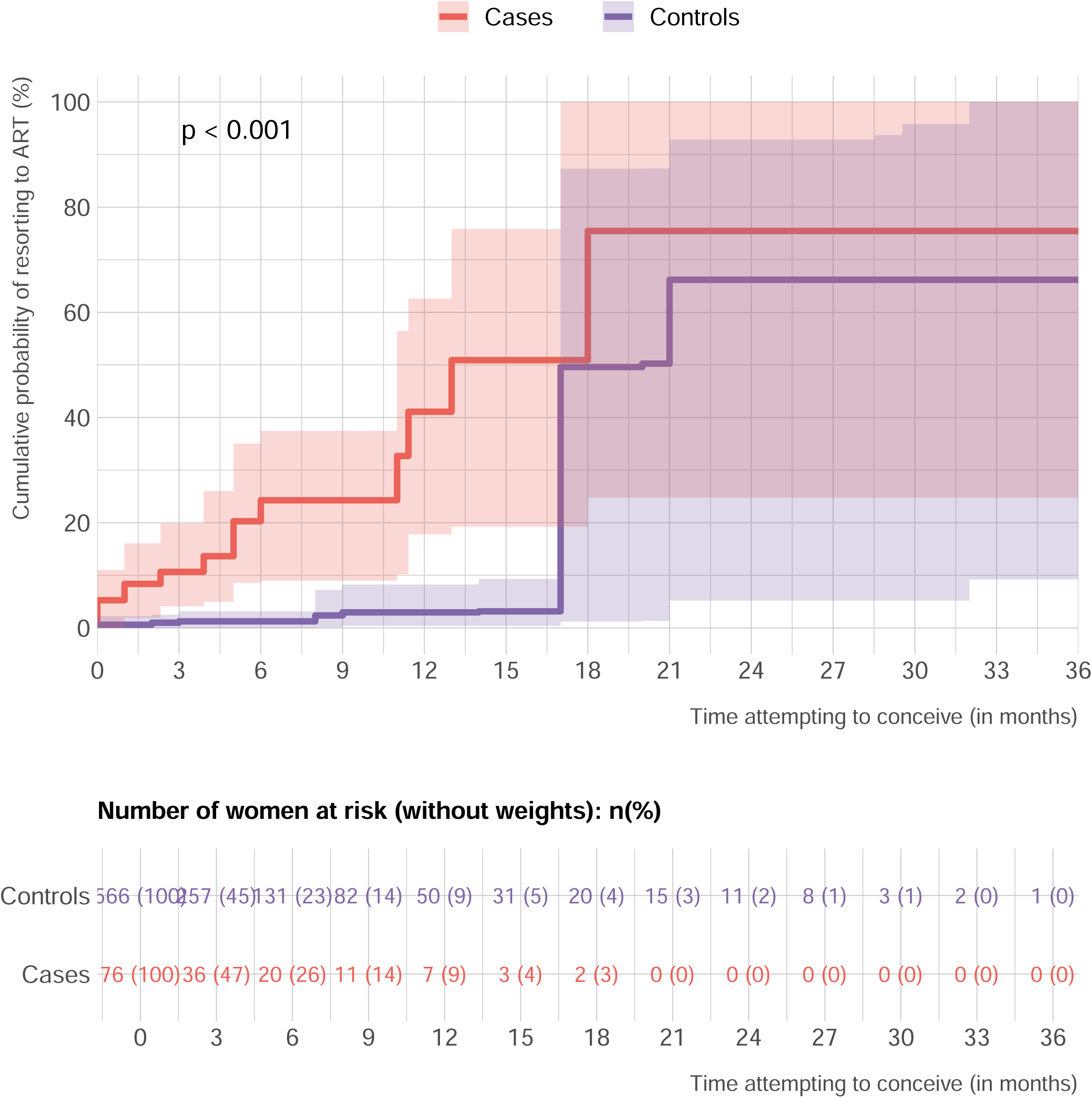
Comparison of time from the first attempts to get pregnant to ART use between cases and controls.

#### Time-to-pregnancy in the subpopulation of women trying to conceive spontaneously

Analyses on the 61 cases and 541 controls who attempted to get pregnant spontaneously (*i.e.* without resorting to ART or using cryopreserved material) showed no significant difference in time-to-pregnancy between cases and controls. The median time-to-pregnancy was 3 months for both cases (95% CI, 2.0 to 3.9 months) and controls (95%CI, 1.5 to 5.0 months), *p*=0.85 (Fig. 4A). One year after the women had started trying to conceive, the cumulative incidence of pregnancy was 82.2% (95% CI, 67.7 to 95.7%) for cases and 82.5% (95% CI, 70.2 to 91.9%) for controls. Two years after they began trying to conceive, the cumulative incidence of pregnancy reached 91.1% (95% CI, 76.1 to 100.0) for cases and 89.4% (95% CI, 77.1 to 99.0) for controls.

**Fig. 4:**
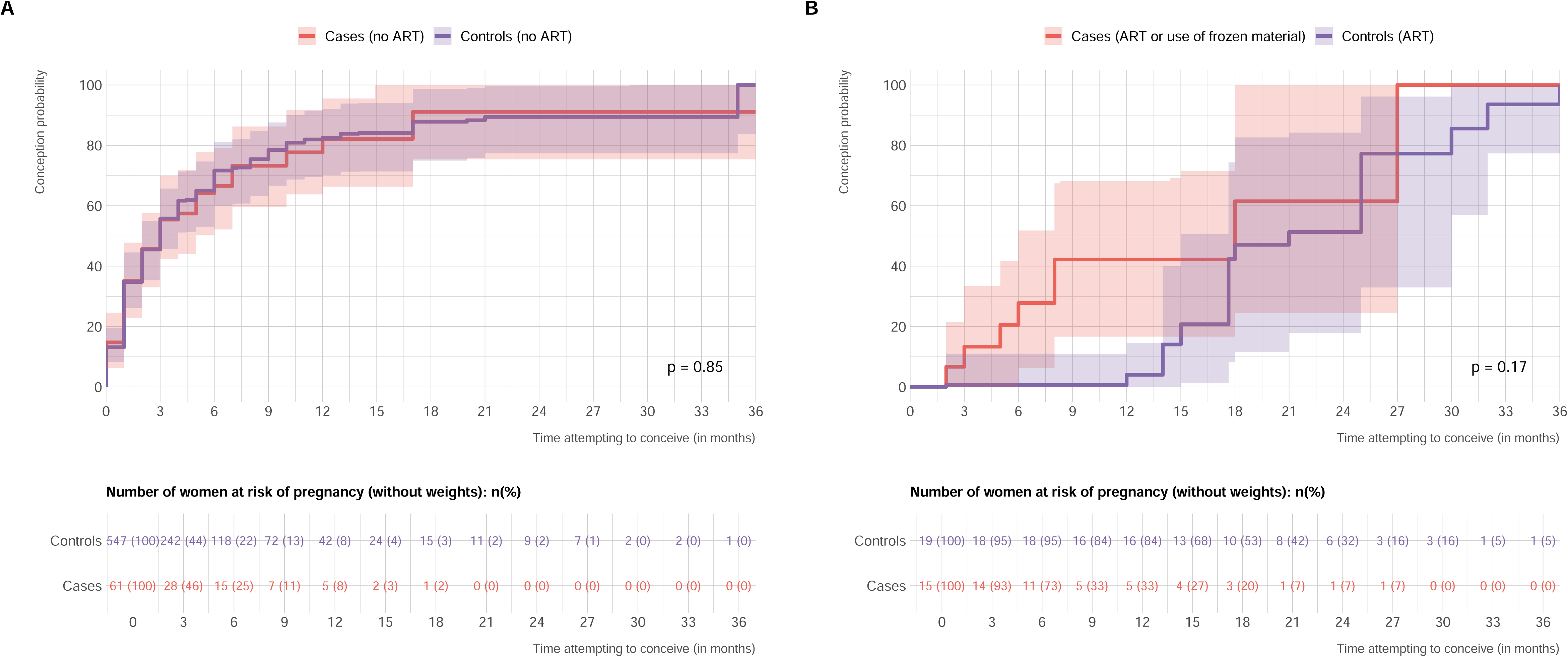
Time-to-pregnancy comparison between cases and controls. **A**: Time-to-pregnancy for women attempting to get pregnant naturally. **B**: Time-to-pregnancy for women using ART, including cryopreserved material.

#### Time-to-pregnancy in the subpopulation of women attempting to get pregnant through the use of ART or cryopreserved material

Analyses performed on the 15 cases and 25 controls who used ART or cryopreserved material showed no significant difference in time-to-pregnancy between cases and controls (Fig. 4B). The median time-to-pregnancy was 18 months for cases (95% CI, 6 to 27 months) and 21 months for controls (95% CI, 15 to 30 months, *p*=0.17). Three cases became pregnant after the use of cryopreserved material (embryos *n*=2, oocyte, *n*=1).

### Factors associated with pregnancy success

Univariable and multivariable analyses identified age at attempt to get pregnant, parity at inclusion, menstrual cycle regularity, BMI, and ART use as independent predictors of time-to-pregnancy, whereas case or control status was not predictive (Table 2).

**Table 2:**
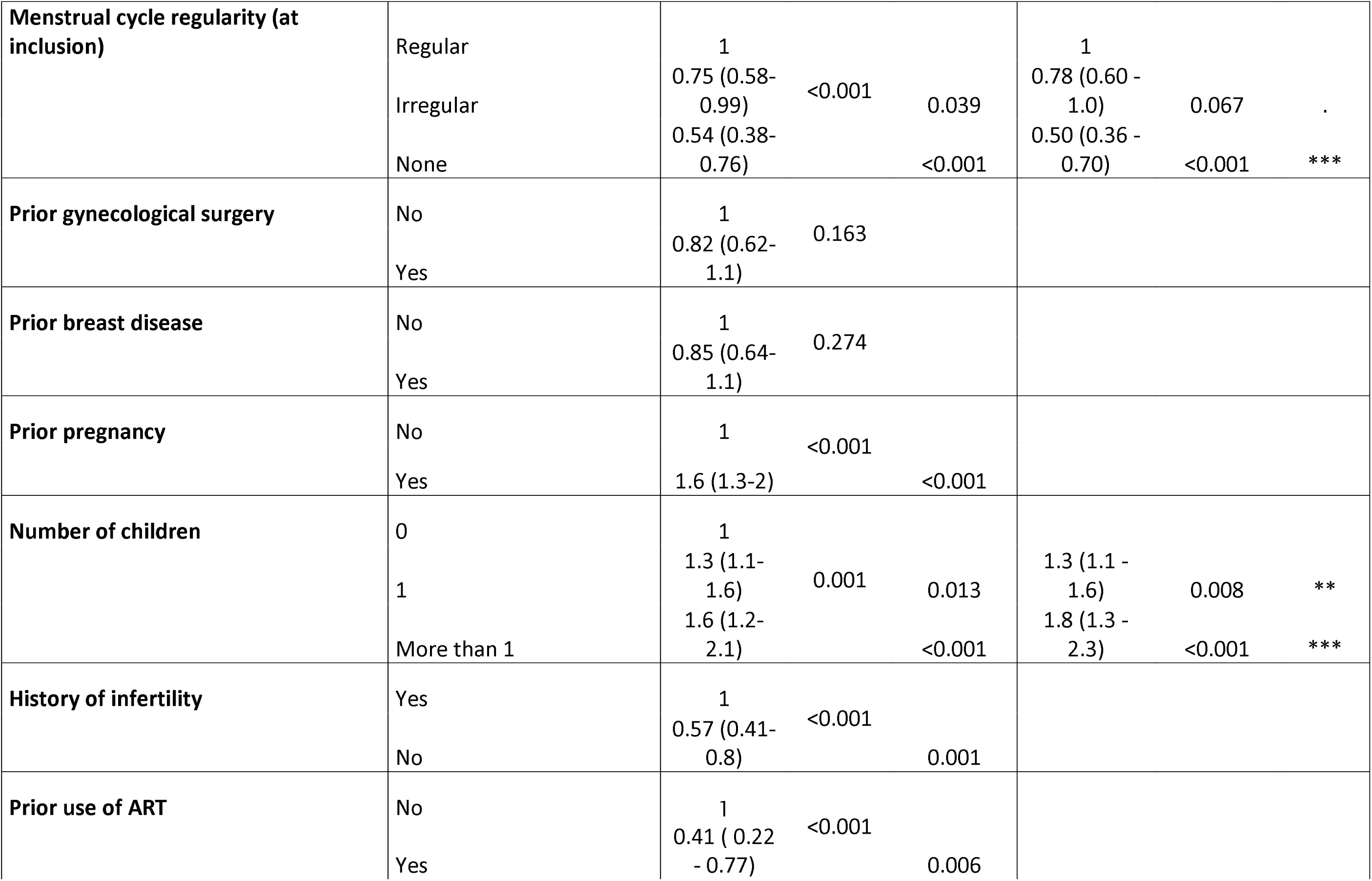
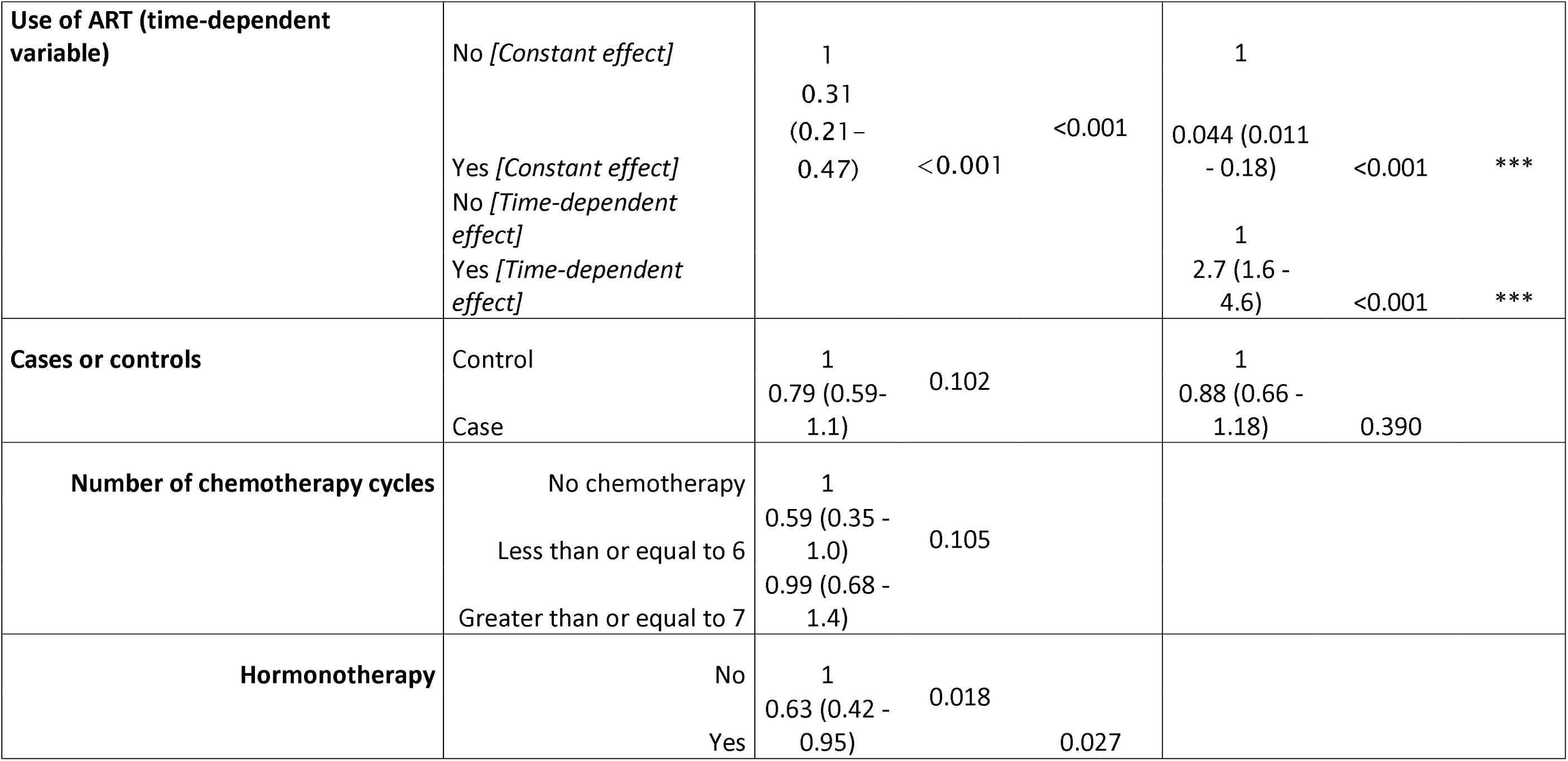
Univariable and multivariable analyses of the factors associated with time-to-pregnancy. The *p*-values for the models refer to comparisons between each class and the reference, and the overall *p*-values from the global log-rank test.

When considered as a binary variable (yes *versus* no), the use of ART methods was significantly associated with a lower probability of becoming pregnant (*p*<0.001). However, this association was variable over time (Fig. S5): ART use was associated with a lower probability of a rapid pregnancy, but the likelihood of pregnancy then increased over time. These findings applied to both BC patients and controls.

### Comparison of pregnancy outcomes between cases and controls

Pregnancy outcomes did not differ significantly between cases and controls (*p* = 0.35). The most frequent outcomes were full-term pregnancies (80.0% for cases *versus* 68.5% for controls) and miscarriages (8.0% *versus* 17.4%, respectively) (FigS6). Cesarean section rates did not differ significantly between cases and controls (24.0% *versus* 12.9%, respectively, *p*=0.11; Fig. S7A). Similarly, there were no significant differences between the groups in terms of the rates of delivery complications (6.0% *versus* 11.1%, respectively, *p*=0.59; Fig. S7B) and rates of malformations in the offspring (4.0% *versus* 2.5%, respectively, *p*=0.82; Fig. S7C).

## Discussion

In this prospective study, we assessed fertility in women with a history of BC and matched controls from the general population. We found no significant difference in time-to-pregnancy in women trying to get pregnant between the two groups.

This finding is unprecedented, as most studies in this field have focused on chemotherapy-induced amenorrhea, fertility preservation options, or fertility biomarkers before and after chemotherapy, but to the best of our knowledge, none has assessed pregnancy rates in patients with a history of BC relative to the general population. Our findings suggest that the probability of pregnancy after BC is not impaired relative to the general population.

Over the last decade, there have been significant advances in fertility preservation techniques, including ovarian tissue cryopreservation, oocyte and embryo freezing, and the use of gonadotropin-releasing hormone analogs. The rapid adoption of fertility preservation techniques was driven by recognition of the potential gonadotoxic effects of cancer treatments and a growing desire of young women to retain their reproductive potential after cancer treatment. The perceived risk and the availability of fertility preservation options led to the widespread use of these techniques, despite the lack of robust, direct data linking cancer treatments to infertility. Endorsement by professional societies and incorporation into clinical practice guidelines have normalized the use of fertility preservation, even in the absence of real data for infertility rates.

In parallel, evidence has emerged to suggest that infertility after BC concerns only a minority of patients. In the Young Women’s Breast Cancer multicenter prospective cohort study, 1026 patients with prior BC enrolled from 12 academic and community sites in the United States from 2006 through 2016 were studied; 69.2% of the women wishing to conceive in this study became pregnant within five years after diagnosis ^24^. In a retrospective French cohort, 133 patients had 197 pregnancies following BC, and the median time-to-pregnancy was 4.3 months^35^. We also analyzed the data available at inclusion for the 517 patients in the cohort studied here with a history of prior BC. Between BC diagnosis and inclusion in the study, 133 pregnancies had occurred in 85 patients, and the median time-to-pregnancy was 3 months ^26^. In the POSITIVE study ^25^, for the 497 patients with a history of prior BC who interrupted endocrine therapy to try to get pregnant and for whom pregnancy status data were available, 74.0% reported falling pregnant at least once during the trial. Notably, premenopausal patients aged 40-42 years had pregnancy rates of almost 50%, providing clear evidence that infertility rates are not particularly high after BC in women from an age class in which pregnancy rates would be expected to be lower.

In our study, women with prior BC were much more likely to use ART or cryopreserved material than controls. However, regardless of whether the pregnancy was spontaneous or resulted from the use of ART or cryopreserved material, time-to-pregnancy was similar in the case and control groups. The rates of ART use in this cohort are lower than those reported in the POSITIVE study, in which 43% of patients used ART. These higher rates could be explained by (i) the greater age of the women in the POSITIVE trial (median age of 37 years *versus* 35.0 years at the start of the attempt to become pregnant in this study); (ii) the study design of the POSITIVE trial, in which the maximum expected duration of the interruption of endocrine therapy — including pregnancy and breastfeeding — was two years after inclusion in the study. Women may therefore have felt obliged to use ART if a spontaneous pregnancy did not occur rapidly.

In our study, the use of ART methods was associated with a lower likelihood of rapid conception, by pregnancy rates gradually increased, reaching those achieved by natural methods. In this cohort, the use of cryopreserved material was rare (8 patients), leading to only three pregnancies. These findings are consistent with those of a recent real-life study ^36^ evaluating the long-term results of 844 fertility preservation procedures, which showed that cryopreserved material was used in only 68 and resulted in only eight live births.

Our study, which had a prospective longitudinal design, is the first to compare pregnancy probability in patients surviving BC with that in the general population. Due to the possibility of residual bias in this comparison, we used state-of-the-art IPTW adjustment methodology to adjust for confounding factors. One of the potential limitations of our study is the recruitment bias inherent in the use of the Seintinelles* network, which is based on voluntary participation, resulting in the enrollment of mostly highly motivated, well-informed women, which could limit the generalizability of our findings. However, both cases and controls were recruited through the same network, so such biases were probably consistent across the two groups.

This study provides significant insight into the fertility outcomes of BC survivors. By showing that the probability of pregnancy in women trying to get pregnant after BC is not significantly different from that in women without BC, it challenges the long-held belief that BC treatments significantly impair probability of pregnancy and highlights the potential for spontaneous pregnancies in patients who have survived BC. From a clinical perspective, these results highlight the importance of comprehensive oncofertility counseling for young patients with BC. Healthcare providers should inform patients not only about fertility preservation options but also about the high likelihood of achieving pregnancy spontaneously after treatment. This balanced approach can help guide informed decisions regarding fertility preservation methods, considering both their benefits and limitations, and emphasizing the natural fecundity of BC survivors.

Once patients who have had BC decide to try to become pregnant, they could be advised to start by trying to conceive naturally rather than resorting to ART methods directly, as natural conception initially outperforms ART. Only after several months of unsuccessful attempts should patients be advised to use ART methods. The precise time cutoff for referral to fertility specialists is unclear, but probably lies at about 6 to12 months, depending on patient age and prior medical history.

These perspectives can foster hope and provide young women faced with BC who hope to have children with realistic expectations, ultimately improving their overall well-being and quality of life after treatment.

## Data Availability

All data produced in the present study are available upon reasonable request to the authors

---

**Fig. S1:**
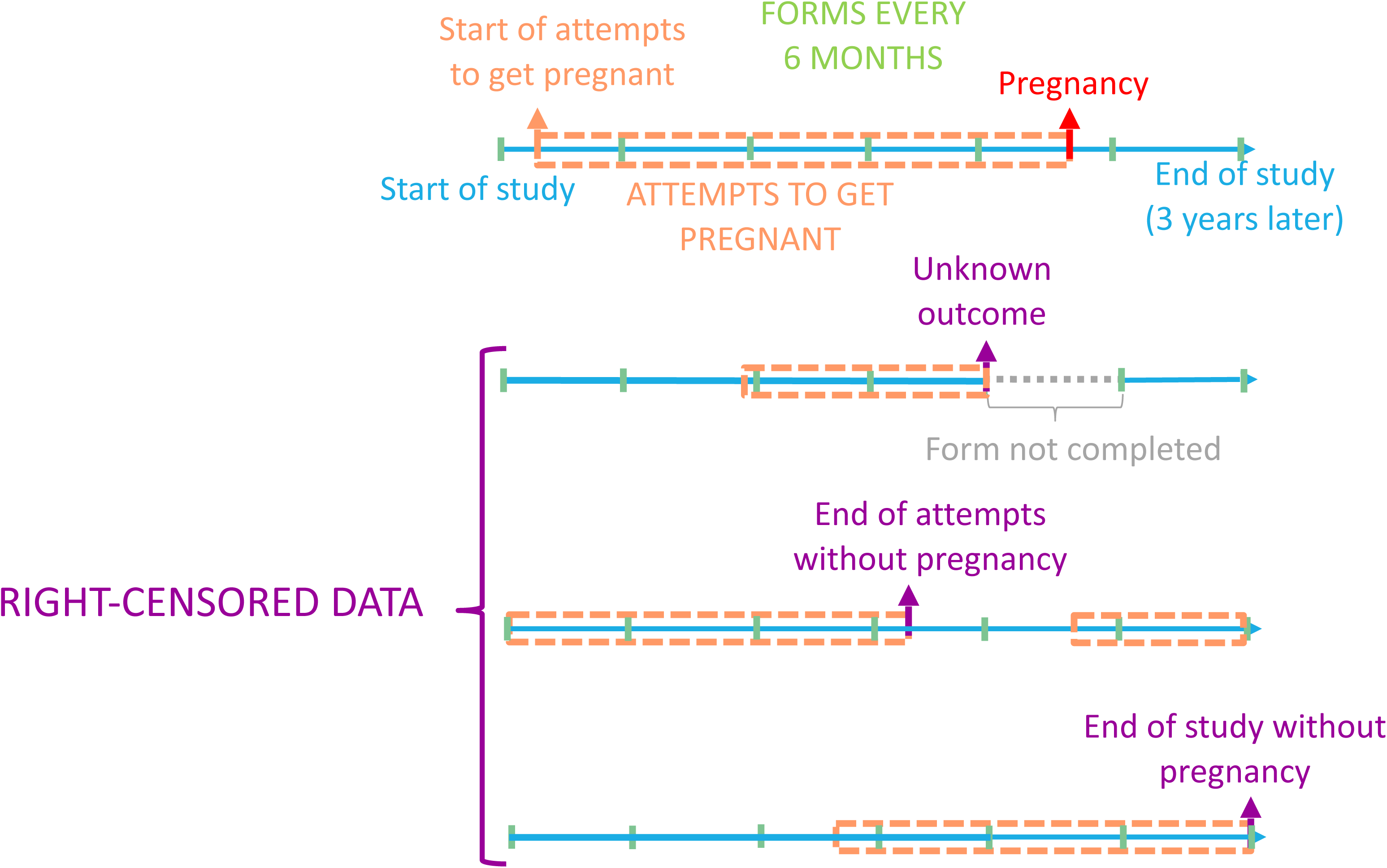
Timelines from the start of attempts to get pregnant to pregnancy or right-censorship.

**Fig. S2:**
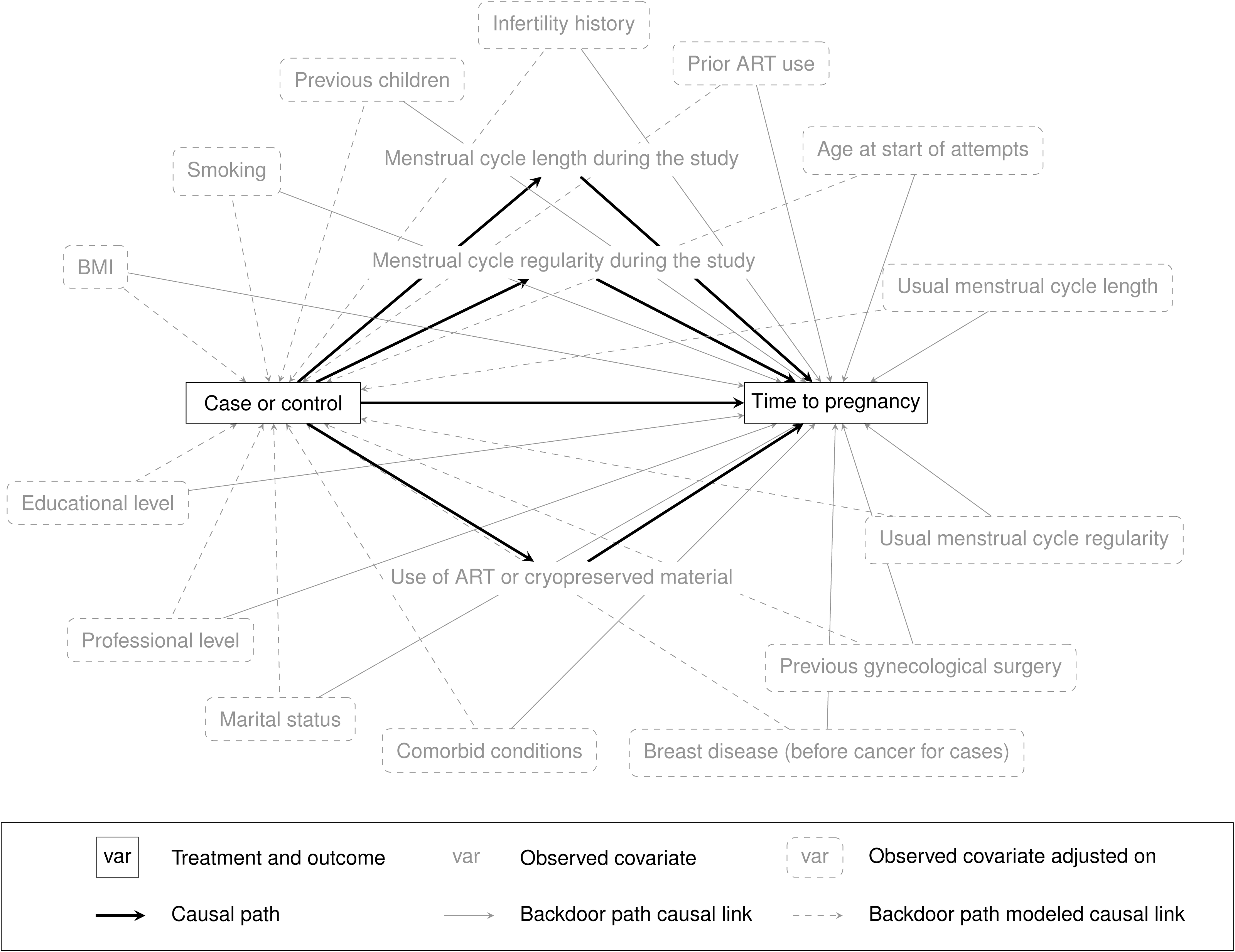
Directed acyclic graph (DAG) clarifying the assumptions underlying the adjustment process. The causal pathway of interest is depicted by bold arrows and involves menstrual cycle length, menstrual regularity during the study, and the use of ART methods or cryopreserved materials. The solid gray lines denote the presumed causal pathways between the 14 covariates and time-to-pregnancy. The dashed lines, while not necessarily corresponding to genuine causal pathways, are treated as such in the analysis due to covariate imbalances between cases and controls resulting from the enrollment methods used.

**Fig. S3:**
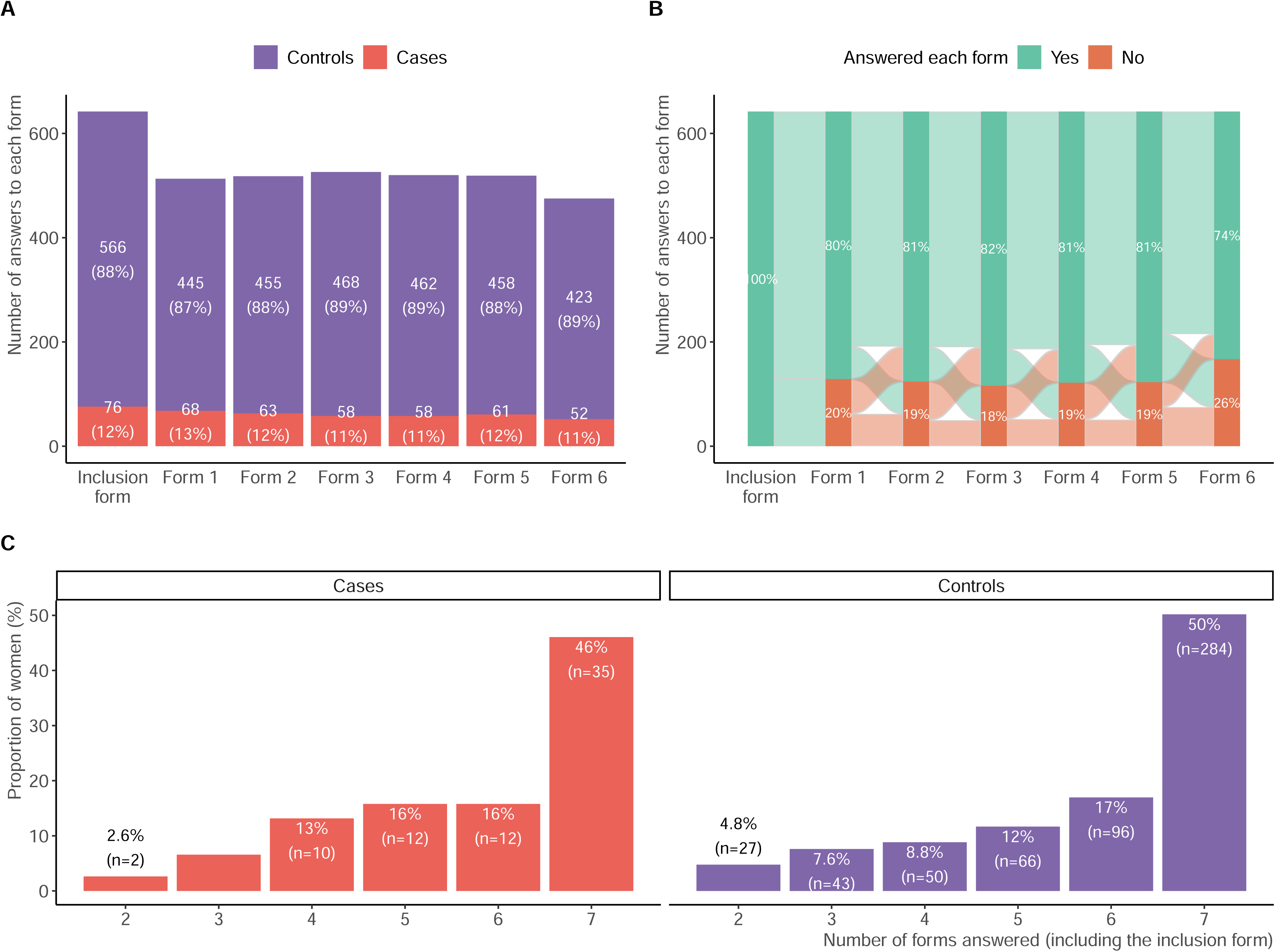
Response rates over the three-year study period. **A**: Proportions of cases and controls completing each form from inclusion to the sixth follow-up form. **B**: Sankey plot representing the successive response rates for each follow-up form. **C**: Total number of forms completed by cases and controls. Each participant selected for analysis completed at least two forms (the inclusion form and one follow-up form).

**Fig. S4:**
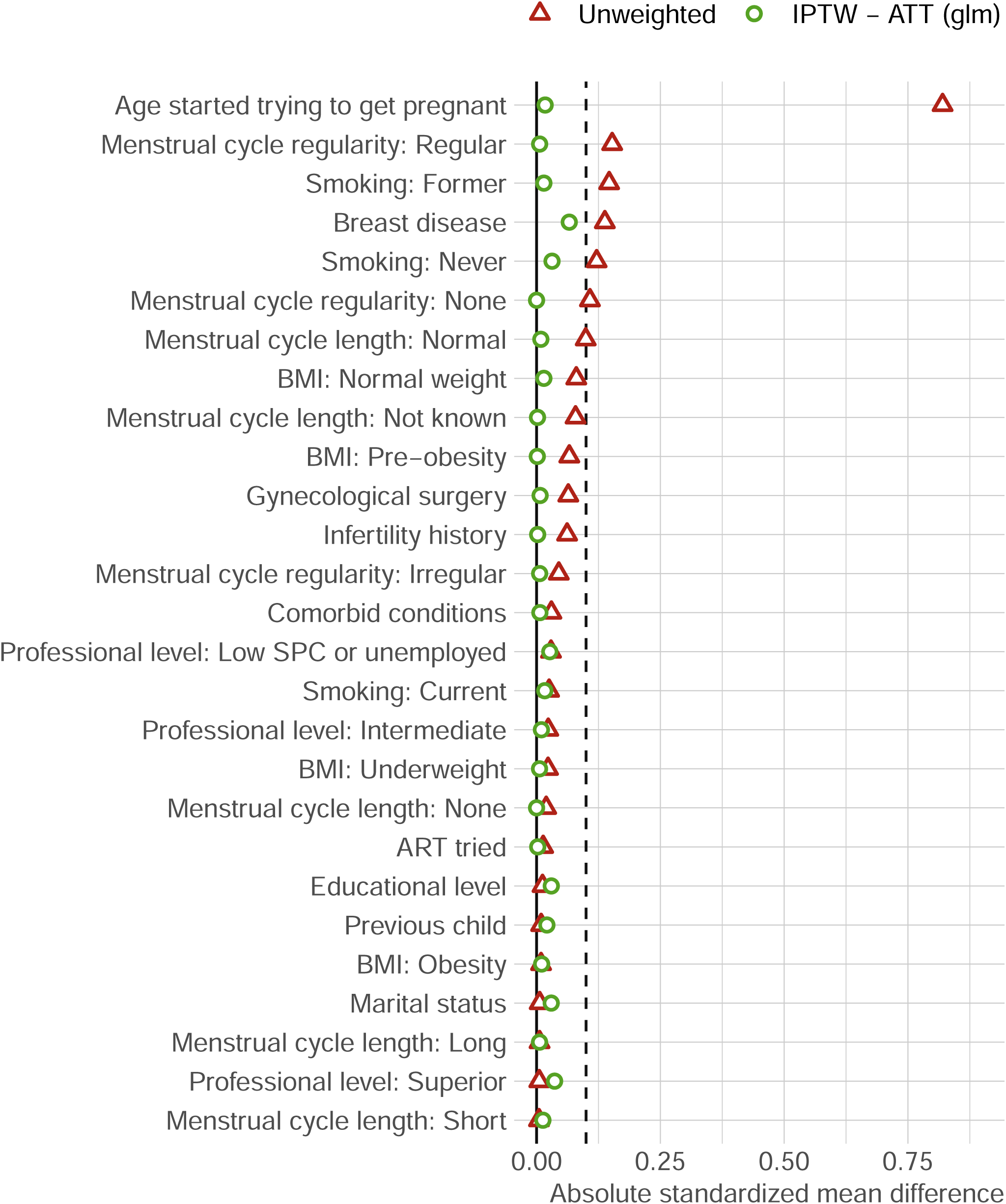
Balance check for the 14 covariates identified as potential confounders for evaluations of the impact of breast cancer treatment on fertility. The standardized mean difference (SMD) evaluates the balance between the values of a given variable in cases and controls. SMD values close to 0 indicate a better balance. Triangles (in red) represent the SMD for unweighted data, showing the imbalance in the covariate distribution between cases and controls before the application of IPTW. Circles (in green) represent the SMD after the application of IPTW ATT weights to controls. The dashed vertical line at 0.1 indicates the threshold for an acceptable balance. Comparisons were performed for 14 covariates both before and after adjustment, to ensure that these covariates did not differ significantly between cases and controls. Abbreviations: IPTW: inverse probability of treatment weighting, ATT: average treatment on the treated.

**Fig. S5:**
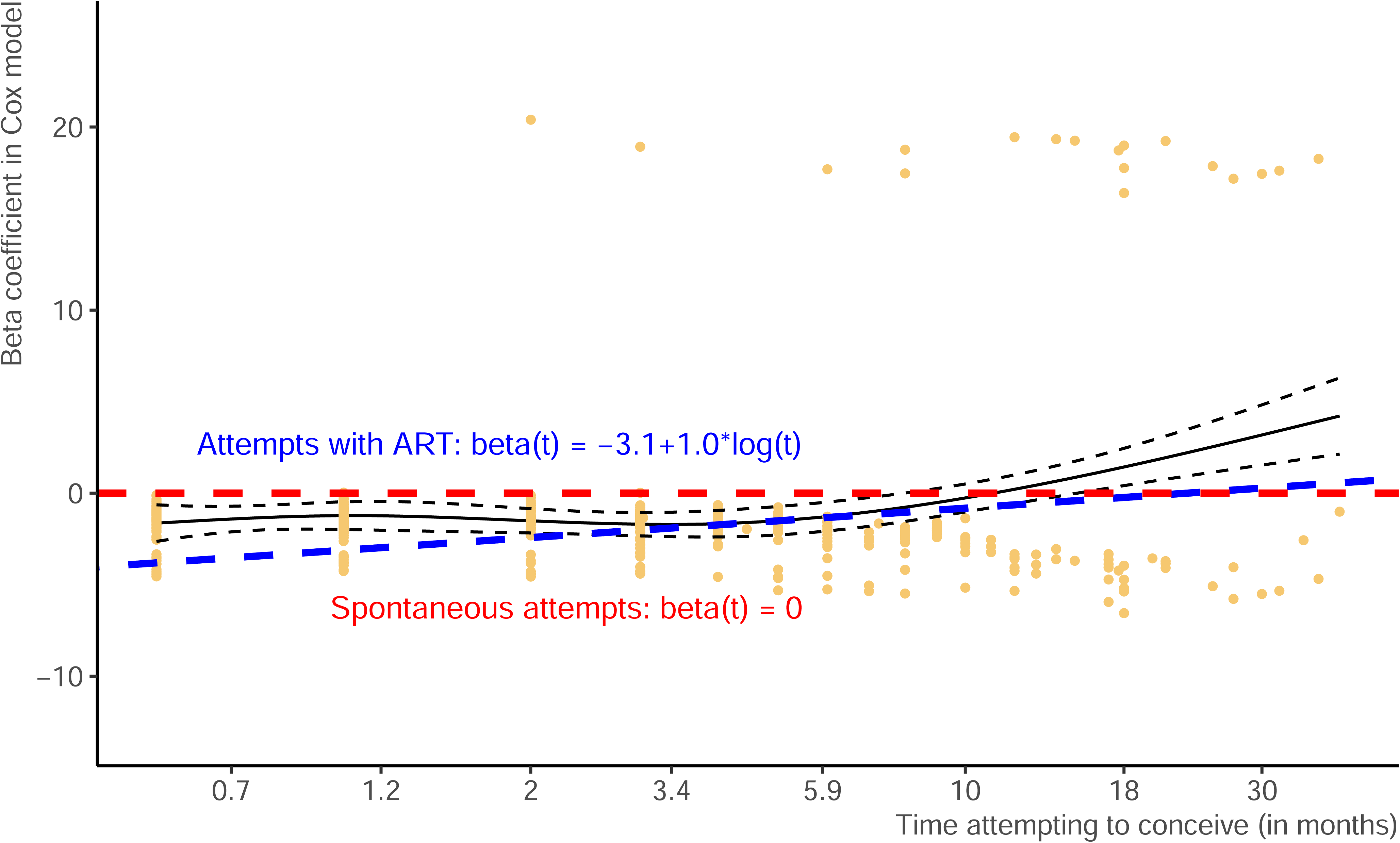
Cox proportional hazards model analyzing the relationship between time-to-pregnancy (TTP) and attempts to become pregnant. The solid blue line corresponds to spontaneous attempts to become pregnant (beta coefficient: 0), whereas the dashed red line corresponds to the use of assisted reproductive technology (ART), with a beta coefficient that changes over time. The *x*-axis represents the time spent attempting to become pregnant (in months), whereas the *y*-axis represents the beta coefficient. The dashed lines represent confidence intervals for the fitted models. The figure highlights the initial lower probability of pregnancy in women using ART, but the probability of pregnancy increases over time in these women, with the curves gradually converging, revealing similar success rates for both methods after a certain period.

**Fig. S6:**
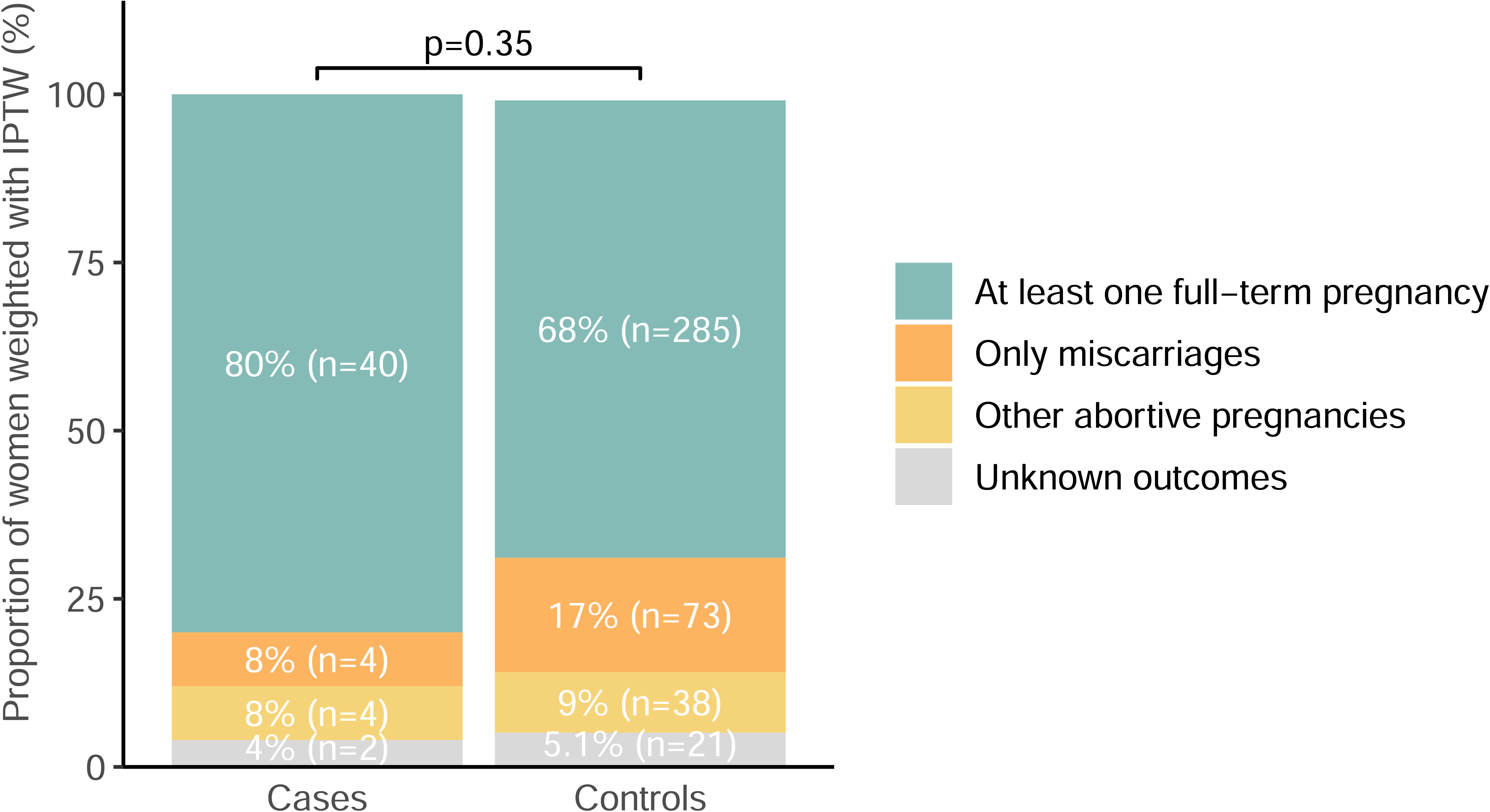
Pregnancy outcomes for cases and controls. Outcomes include full-term pregnancies, miscarriages, and other abortive pregnancies (elective abortion, abortion for medical reasons, and ectopic pregnancy). “Unknown outcomes” are pregnancies for which the outcome was not reported, often due to subsequent forms not being completed or pregnancies that had not yet been completed at the time of the last questionnaire. Proportions were calculated by applying IPTW ATT weights to the control group to match the distribution of confounding variables in the cases. The raw numbers were then determined by taking the closest integer to the product of the weighted proportion and the actual number in each category. A *p*-value was obtained in Fisher’s exact test. Abbreviations: IPTW: inverse probability of treatment weighting, ATT: average treatment on the treated.

**Fig. S7:**
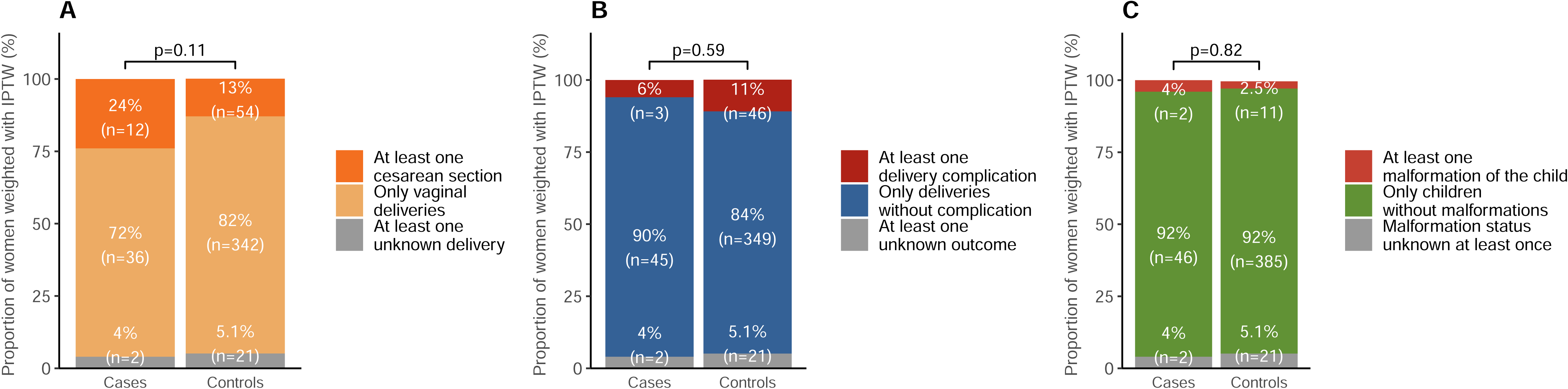
Types of delivery, complications and child malformations for cases and controls. Proportions were calculated by applying IPTW ATT weights to the control group to match the distribution of confounding variables in the cases. The raw numbers were then determined by taking the closest integer to the product of the weighted proportion and the actual number in each category. The *p*-values were obtained in Fisher’s exact test. **A**: Proportions of women with at least one cesarean section, as opposed to vaginal deliveries. **B**: Proportions of women with at least one delivery complication. **C**: Proportions of women for whom the child presented at least one malformation. Abbreviations: IPTW: inverse probability of treatment weighting, ATT: average treatment on the treated.

**Table S1:**
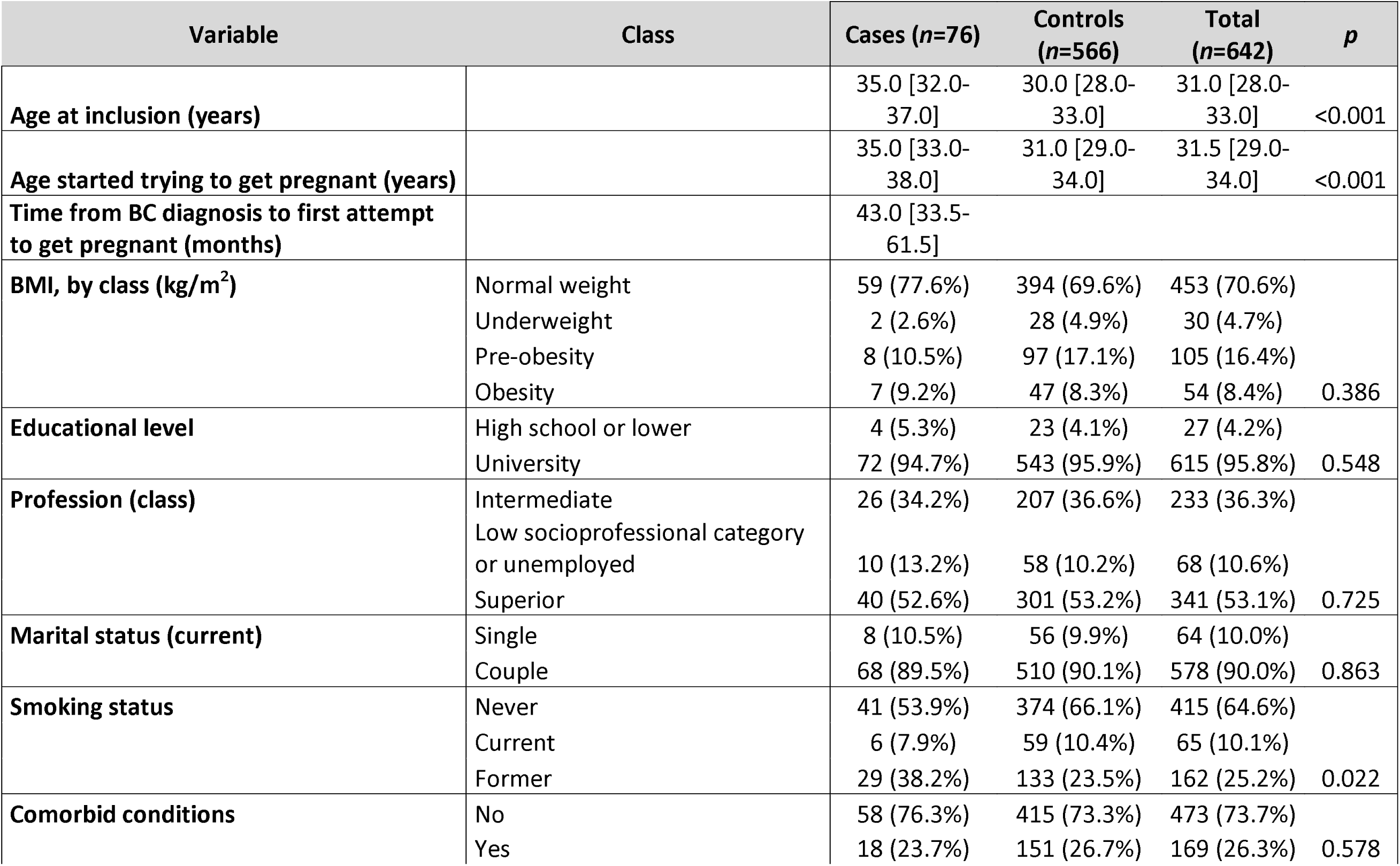
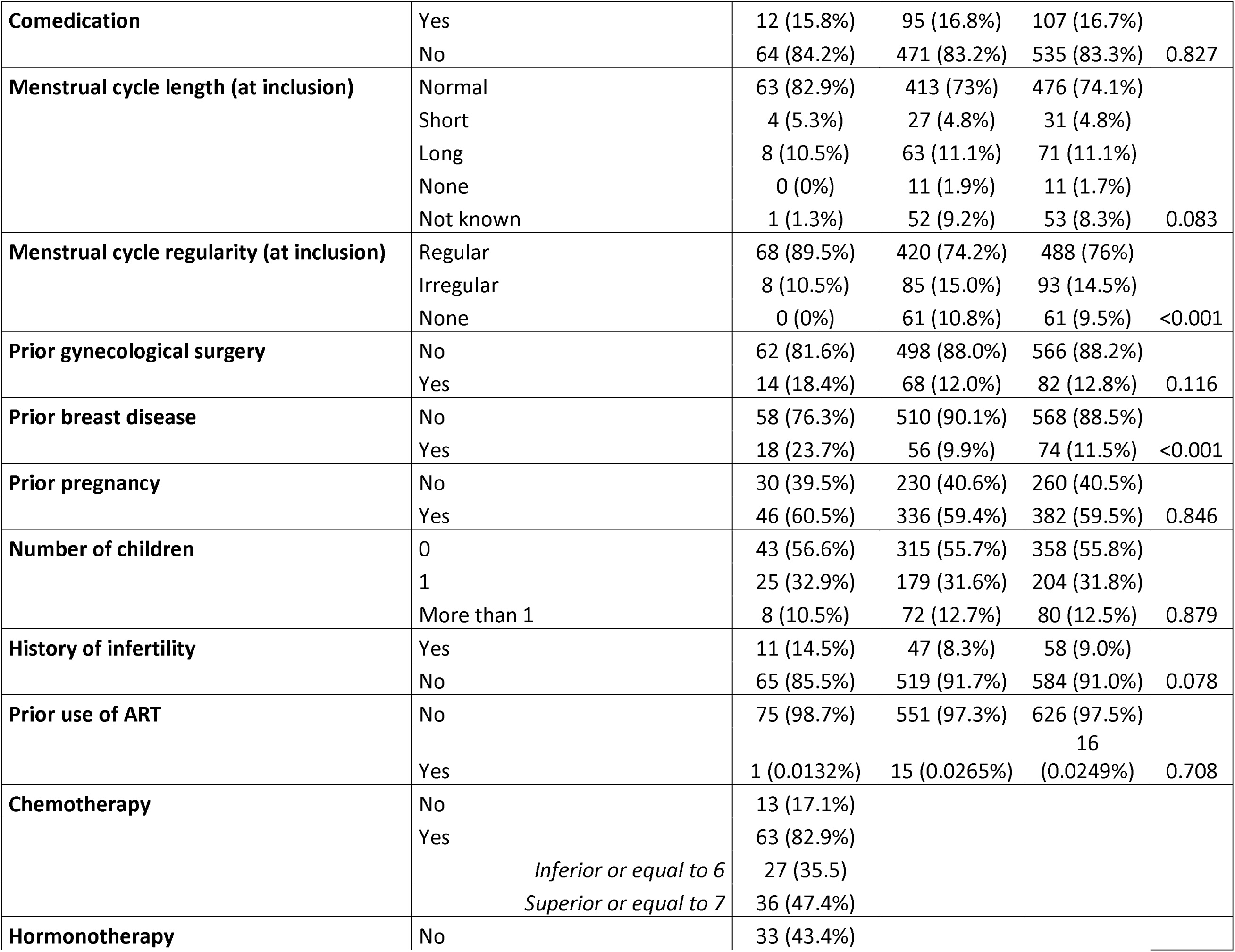
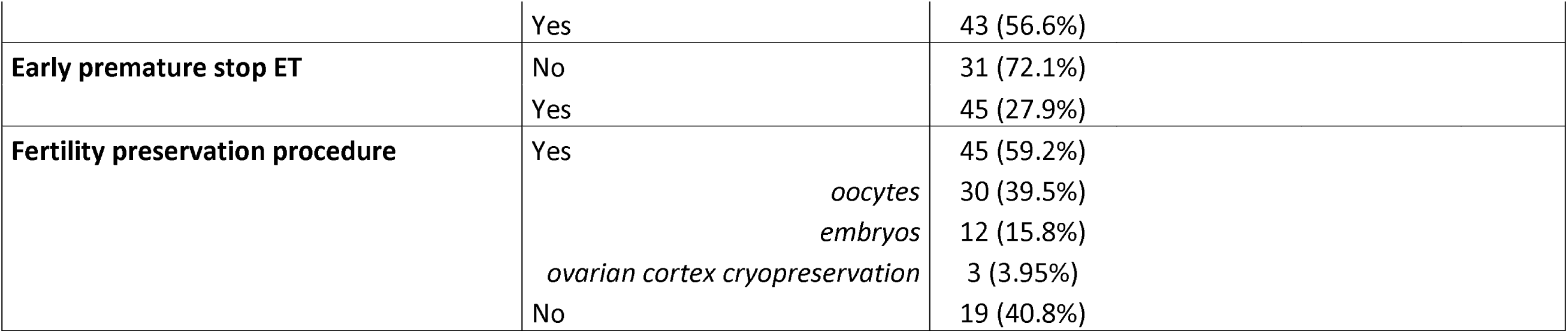
Characteristics of the study participants before adjustment. This table presents the demographic and clinical characteristics of the study participants, comparing women with (cases) and without (controls) a history of breast cancer.

## Authors’ contributions

Conceptualization: ASH, AC, CRJ, CB, LS, ME, GPB, LMM, GJ, FC, FR

Data curation: ASH, AC, CS, CB

Formal analysis: AC

Funding acquisition: ASH, FR

Investigation: ASH, LS, AB, WJ, ME, SG, GPB, LMM, GJ, FC, FR

Methodology: ASH, AC, ED, FJ, BA

Project administration: ASH, LS, LD, AT, RB, GJ, FC

Software: ASH, ED

Resources: ASH, CRJ, CB, LS, LD, AT, CD, LSo, GJ, FC, FR

Supervision: ASH, IRC, AG, BA, GJ, FR

Validation: ASH, CRJ, AB, WJ, ME, SG, AT, PG, BA, GJ, FR

Visualization: ASH, AC, ED, FJ, PG

Writing – original draft: ASH, AC

Writing – review & editing: ASH, AC, CRJ, CB, IRC, LD, ED, AB, WJ, ME, SG, GPB, LMM, AG, PG, RB, CD, LSo, GJ, FC, FR

## Information on author access to data

Data accessible upon reasonable request by contacting the corresponding author.

## An explanation of the role of funder(s)/sponsor(s)

The funder was not involved in study design, or in the collection, analysis and interpretation of data, the writing of this article or the decision to submit it for publication.

**The authors thank all the study participants from the Seintinelles Research Network.**

