## supplemental material for "A prospective exposed-unexposed cohort study comparing time-to-pregnancy in patients with previous breast cancer and controls from a collaborative research network: results after three years of follow-up"

**Materials and methods**

***Statistical analyses***

The study population is described in terms of frequencies for qualitative variables, or means, medians and associated ranges for quantitative variables. No data were missing for the continuous variables, and for the qualitative variables, a category “Unknown” was created when needed. For continuous variables, Wilcoxon-Mann-Whitney tests were used for comparisons of groups containing fewer than 30 patients or for comparisons in which one group showed signs of non-normality (indicated by a Shapiro test *p*-value of 0.05 or less). Otherwise, we used the Welch *t-*test. The association between categorical variables was assessed in Fisher's exact tests if at least one category included fewer than five patients or the Chi-squared test of independence without continuity correction otherwise.

***Censorship***

Women were subject to right-censoring under the following conditions: i) not completing a form during the period in which she was trying to get pregnant, ii) decision not to continue trying to get present, for personal reasons, iii) end of the three-year study period reached without the occurrence of a pregnancy (Fig. S1). These three conditions were considered uninformative and independent of the likelihood of conception, justifying the censoring. Relapse should have been treated as a competing risk, but none of the cases experienced a relapse during the period of the study in which they were trying to get pregnant.

***Confounding factors and statistical handling***

We identified 14 covariates associated with conception that were not equally distributed between cases and controls and might act as confounders in our dataset: age at which the woman first started trying to get pregnant, nulliparity, professional and educational levels, marital status, body mass index (BMI), smoking status, infertility history, previous use of ART methods, menstrual cycle length and regularity, prior gynecological surgery, prior breast diseases (before cancer for cases) and comorbid conditions. We identified these covariates based on clinical information, and the results obtained with univariate and multivariate Cox models. The directed acyclic graph (DAG) (Fig. S2) clarifies the assumptions underlying the adjustment process.

The propensity score method involves assigning a weight to each control such that the weighted control group has the same overall distribution of each confounding variable as the case group. The weights remained within a reasonable range and did not require stabilization or trimming. We assessed covariate balance after weighting by calculating the standardized mean difference (SMD) and the Kolmogorov-Smirnov (KS) statistic, with balance thresholds of 0.1 and 0.05, respectively ^34,35^. The positivity assumption was checked variable by variable, and by looking at propensity score overlap. The confidence intervals for the Kaplan-Meier curves were obtained by computing 1000 bootstraps ^36^. For the bar charts, proportions were calculated by applying weights to the control group to match the distribution of confounding variables to that in the cases. The raw numbers were then determined by taking the closest integer to the product of the weighted proportion and the actual number in each category. This approach ensures that the bar charts accurately reflect the adjusted comparisons between the groups.

***Cox proportional hazards model***

The model was fitted by the “add1” method, according to which variables are added until no further improvement in the model fit is observed. This process involves conducting a likelihood ratio test at each step, comparing the likelihood of the model with the added variable and the likelihood of the model without the added variable. The variables “case or control” and “age when started trying to get pregnant” were forced into the model. In this analysis, patients were not weighted by IPTW, to allow the evaluation of several of the variables among the 14 confounders identified for time-to-pregnancy. The model incorporated a time-dependent variable ^38^ and we addressed the violation of the proportional hazards assumption for one variable (identified by testing the correlation of Schoenfeld residuals with time) ^39^ by introducing affine time-dependent coefficients into the model ^40,41^.
